# Actions for fostering cross-disciplinary global health research A qualitative research

**DOI:** 10.1101/2021.09.30.21264294

**Authors:** Yan Ding, Ewan Tomeny, Imelda Bates, the IMPALA Consortium

## Abstract

**Introduction:** The ability to conduct cross-disciplinary research in international collaborations is critical for improving global health. Published evidence on actions to foster cross-disciplinary research comes mainly from high-income countries with its applicability to global health unclear. Our study investigated the actions taken to foster cross-disciplinary research across a North-South global health programme to recommend actions to improve the effectiveness of future cross-disciplinary research programmes in global health.

**Methods:** We used an adapted three-component framework to frame data collection and analyses. Building on a recent literature review, we compared actions used by the programme for planning, implementing and managing cross-disciplinary research to those described in the literature. Data sources included interviews with 31 participants (including researchers, administrators, and collaborators), a review of programme documents, a baseline survey, and observations of interactions at meetings and events. Interview data were coded and analysed using the framework. Narrative summaries were created using thematic synthesis and triangulated by the document review and observations.

**Results:** For cross-disciplinary research to be successful in global health, a shared vision with explicit goals is essential. These goals concern knowledge integration supported by regular communication, orientation about involved disciplines, and equitable partnerships across institutions and individuals. Fostering cross-disciplinary global health research needs a significant time investment from researchers, support staff, and programme leaders. Indicators for tracking cross-disciplinary working should be agreed at the outset and monitored throughout.

**Conclusion:** Cross-disciplinary activities should be managed separately from primary research activities. The three-component framework would be helpful in guiding designing cross-disciplinary programmes.

**Summary Box:** *What is already known?:*

- Global health can be advanced by cross-disciplinary collaboration within and beyond the health sciences.
- Individual researchers, research team leaders, academic institutions and research funders all have roles in making cross-disciplinary research more effective.
- Published evidence on actions to foster cross-disciplinary research comes mainly from high-income countries and its applicability to global health is unclear.

*What are the new findings?:*

- The cross-disciplinary aspects of research programmes need to be actively managed.
- Pre-agreed indicators should be used to plan and track cross-disciplinary research.
- Fostering cross-disciplinary research and managing tensions takes time and explicit continuous discussions.
- Our adapted three-component framework (i.e., planning, implementation and management) is useful for collecting and analysing multi-source and multi-perspective data on a cross-disciplinary programme in real-time.

*What do the new findings imply?:*

- Cross-disciplinary activities should be managed and tracked separately from the primary research activities.
- Progress in research planning, implementation, and management should be reviewed against pre-agreed indicators, with troubleshooting conducted accordingly.
- Cross-disciplinary research requires the allocation of more time and funds for active management of research planning than would be required for mono-disciplinary projects.
- The three-component framework would be helpful in guiding the design of cross-disciplinary programmes.

## INTRODUCTION

Bringing together researchers from multiple disciplines can lead to innovation and rapid production and dissemination of cross-disciplinary knowledge to solve complex global health problems.[1, 2] Cross-disciplinary research has been growing globally in popularity among researchers and funders because of its importance in addressing global health challenges.[1, 2] ‘Cross-disciplinary research’ covers three typologies: multi-, inter-, and trans-disciplinary research. In this article we will use the term cross-disciplinary research (CDR) to mean research that combines concepts, methods, and theories drawn from two or more disciplines.[3]

Existing evidence on fostering CDR is fragmented across disciplines,[4, 5] making it difficult to find. There is increasing interest in understanding how to implement effective CDR and in the importance of team dynamics between researchers from disparate disciplines. CDR tends to be more complex than traditional types of research,[6] and presents unique challenges,[1, 3] such as problem definition, positioning in different disciplines[7] and coordination of effort.[8, 9] Our previous literature review found that evidence about how to conduct effective CDR is primarily from high-income countries, and may not apply to CDR in global health, where research is typically conducted through North-South collaborations.[3]

We used the *International Multidisciplinary Programme to Address Lung Health and Tuberculosis in Africa* (IMPALA 2017-21))[10] as a case study to explore and reflect on practical actions for fostering CDR in North-South collaborations. IMPALA aimed to generate knowledge and implementable solutions concerning lung health and tuberculosis. Led by a Global North research institute, IMPALA had 22 international partner organisations from 13 countries, 10 in sub-Saharan Africa.

IMPALA explicitly used multidisciplinary approaches and spanned biology to policy[11]. It involved five research disciplines: *clinical science, social sciences, health systems, health economics*, and *policy/research capacity*. Unusually, to promote fairness and overcome disciplinary hierarchies, the programme was framed around these discipline groups: each group initially received the same amount of funding and was represented on the management team alongside the three consortium directors. Each group had one PhD student and one postdoctoral researcher (Figure 1), with equal training opportunities offered to all early career researchers (ECRs).

**Figure 1:**
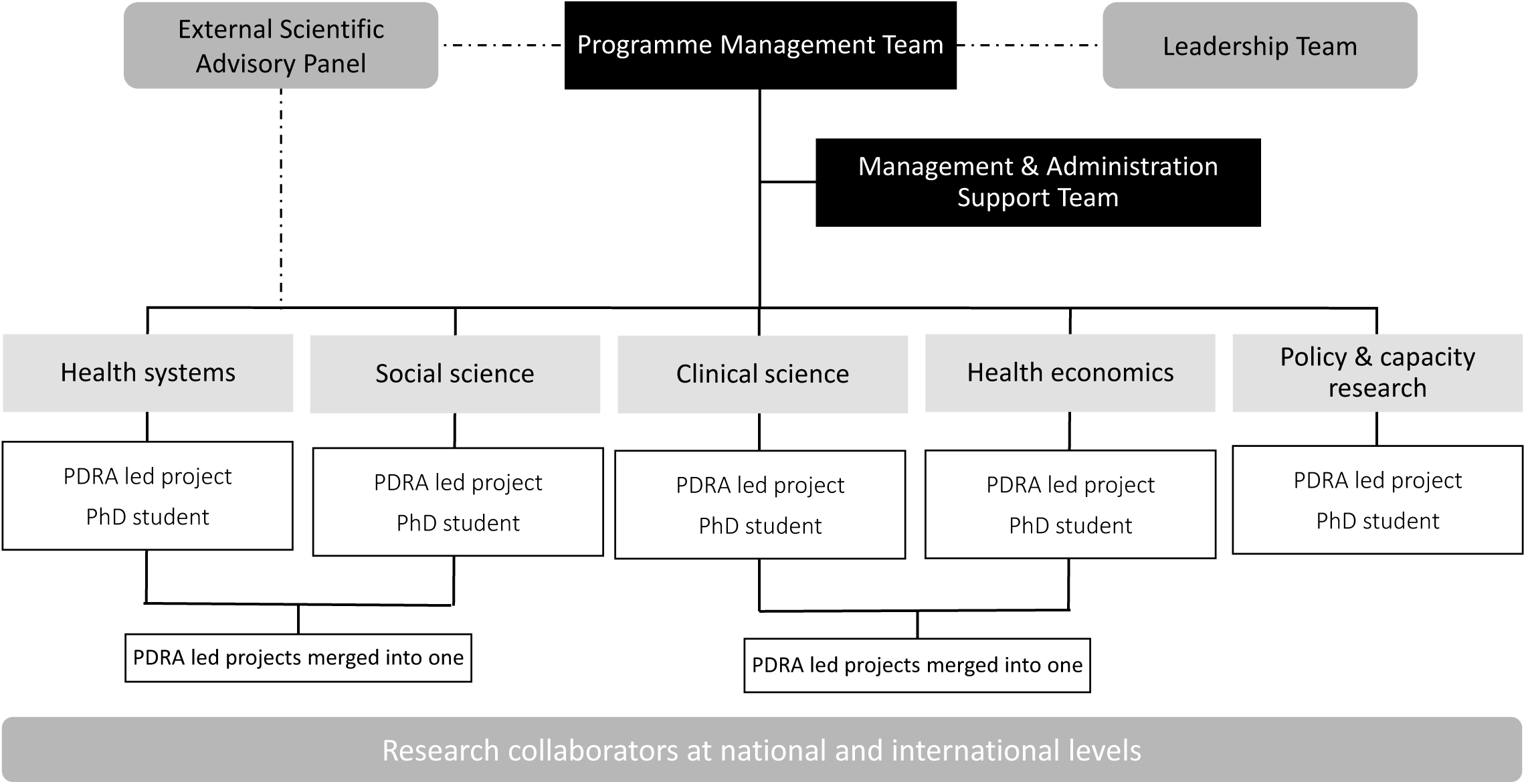
The IMPALA Organogram

Our study has drawn on IMPALA as a whole, and its two embedded projects (hereafter *‘the two projects’*): one combined *clinical sciences* and *health economics*; the other *health systems* and *social sciences*. Our study explores the ‘real life’ actions taken to foster CDR across IMPALA and based upon our findings we recommend actions to improve the effectiveness of future global health CDR programmes.

## METHODS

We adapted a four-phase model of CDR collaborations (Figure 2)[9] which describes objectives within each project phase (i.e., *Development, Conceptualisation, Implementation, Translation*). We combined the *Development* and *Conceptualisation* phases into one ‘Planning’ phase, since global health research activities in these phases are generally integrated.[12] The *Translation* phase was not included because it requires long-term follow-up. Our literature review indicated that leadership and management strongly influence CDR effectiveness,[3] so these were added as a cross-cutting framework component.

**Figure 2:**
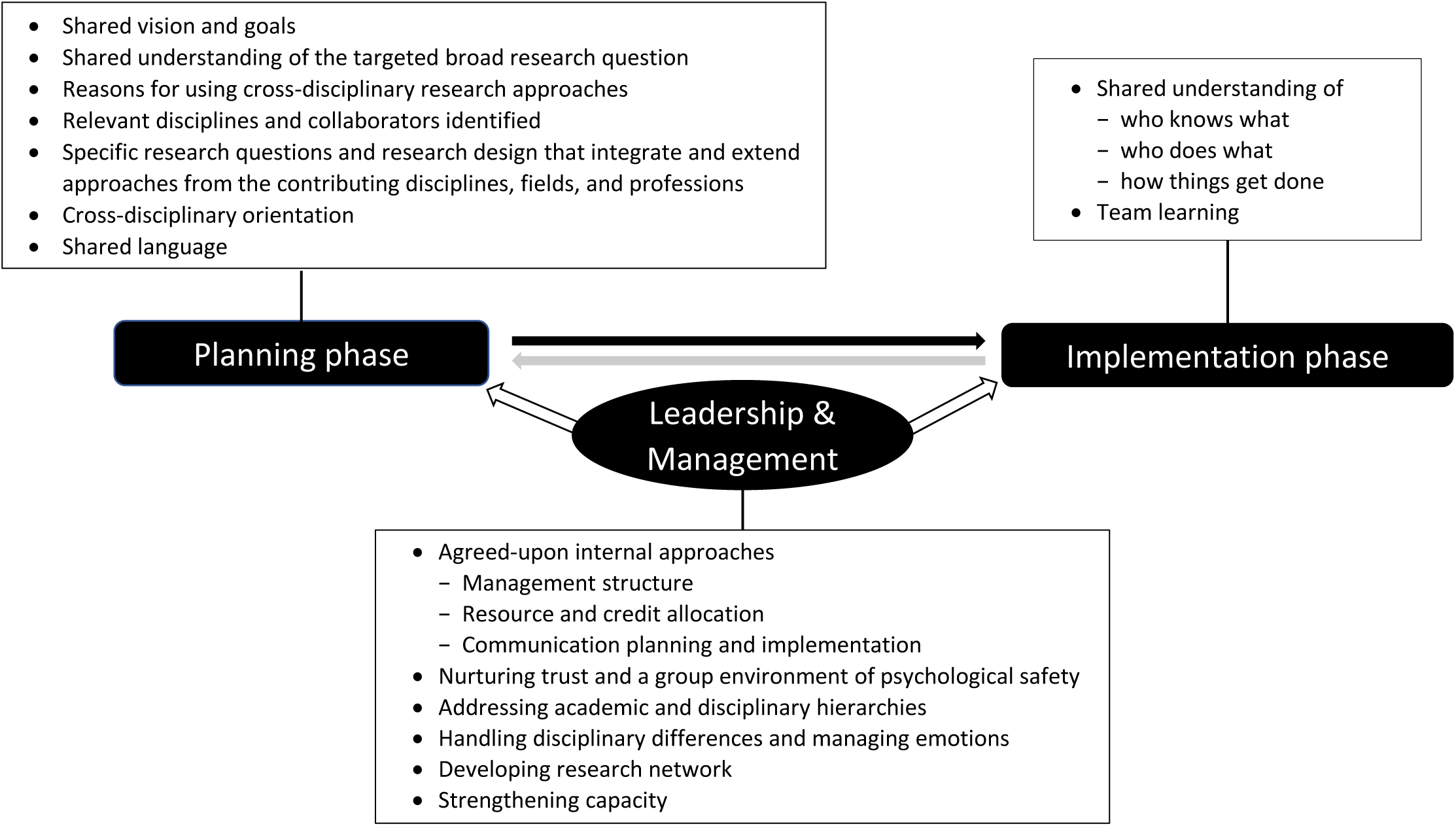
The three-component framework for the cross disciplinary collaborative research process used in this study (adapted from Hall et al., 2012)

### Data collection

Our primary source of data was semi-structured interviews, supplemented by a baseline survey, a document review, and observations of events.

#### Baseline survey

All IMPALA members were invited to complete a baseline survey (May-September 2018). This included individuals from the External Scientific Advisory Panel, Leadership and Management Teams, Administrators, and researchers/policy makers involved in the two projects. The survey collected participants’ personal information, and their experience of, and confidence in, conducting CDR. The survey findings were used to identify potential interviewees and tailor interview guides.

#### Semi-structured interviews

##### Sample selection

The interviews collected data on challenges and practical actions/solutions related to the fostering and conducting CDR. Guided by our baseline survey data 31 primary interviewees were selected using the IMPALA team directory as a sampling frame. Purposive sampling was used to maximise variation in roles, disciplinary backgrounds, career stages, gender, affiliated organisations, and geographical locations[13].

##### Procedure of interviews

Interview questions were based on the adapted framework (Figure 2) with probes informed by our literature review.[3] Interviews were audio recorded and conducted in English, either in-person or virtually. Participants received an information sheet prior to their interview and provided informed consent. For anonymity each interviewee was assigned an ID.

#### Document review

Data were extracted from documents concerning the programme’s: *vision, goals, research questions, design, teams, interactions*, and *outputs*, to understand the context of the programme and its projects, inform interview questions, and cross-check findings from other data-collection methods. Documents included the IMPALA website, concept notes, proposals, minutes/agendas from annual meetings, and quarterly research updates.

#### Observation of events

YD was a participant observer at events involving cross-disciplinary issues including two annual meetings, monthly knowledge exchange meetings, training workshops, and a 4-day field visit to Tanzania. Observation notes were entered in real time into a pre-designed form informed by the literature,[14] comprising sections on: brainstorming the crossing of analytical levels; integration of disciplinary ideas; proposed/actual cross-disciplinary outcomes; information sharing; technical or emotional support; and challenges and setbacks.[14] Observation findings were used for refining interview questions and triangulating interview data.

### Data analysis

Interview data were coded, mapped and analysed using the framework (Figure 2). Narrative summaries were created using thematic synthesis.[15] Findings were triangulated using the document review and observations.

### Research ethics

Ethical approval was provided by Liverpool School of Tropical Medicine’s Research Ethics Committee (Reference: 18-031).

### Patient and public involvement

While the IMPALA programme involved both patients and the public, due to this study’s specific focus on research practice, its design, conduct, and reporting did not involve patients or the public.

## RESULTS

### Interviewee characteristics

Thirty-six interviews with 31 interviewees were conducted each lasting 68-192 minutes. Five individuals were interviewed again after one year to identify changes in CDR in the two projects. 52% of interviewees (16/31) were female, and 16 were based at African organisations from 7/10 partner African countries. Twenty-five (81%) were researchers, comprising 18 senior researchers (i.e., professors, readers, associate professors, senior lecturers) and 7 ECRs (i.e., assistant professors, lecturers, research fellows, post-docs, research assistants, PhD students) (Table 1).

**Table 1:** Interviewees’ characteristics

|  | Items | Option | N |
| --- | --- | --- | --- |
| 1 | Role in IMPALA<br>(Options not mutually exclusive) | A member of the External Scientific Advisory Panel | 2 |
|  |  | A member of the Leadership Team | 4 |
|  |  | A member of the Management Team | 8 |
|  |  | A member of the Management & Administration Support Team | 3 |
|  |  | Other member working across IMPALA projects | 2 |
|  |  | A researcher or policy maker on the two projects; | 15 |
|  |  | (of these those based in Africa) | (8) |
|  |  | A member who was based in Africa, working on IMPALA, but not on the two projects | 5 |
| 2 | Sex | Female | 16 |
|  |  | Male | 15 |
| 3 | Location | African Countries | 16 |
|  |  | United Kingdom | 15 |
| 4 | Primary Disciplinary Background | Medicine & Clinical Sciences | 18 |
|  |  | Humanities & Social Sciences | 10 |
|  |  | Others | 3 |
| 5 | Profession | Researchers/research leaders | 25 |
|  |  | Non-research members | 6 |
| 6 | Academic Rank (of the 25 researchers) | Senior researcher | 18 |
|  |  | Early career researcher | 7 |

### Survey, document reviews and observations

The baseline survey was sent to 66 IMPALA staff, with responses received from 43/56 researchers (77%), and 8/10 non-researchers (80%). Twenty-four events were observed over 20 months (Sept. 18 – Apr. 20) and 49 documents were reviewed (Box 1).

#### Box 1

Internal IMPALA documents used to provide background information for this study

1. The IMPALA website: https://www.lstmed.ac.uk/impala
2. IMPALA Technical Proposal
3. IMPALA Team Directory
4. The concept notes of all the eight research projects sitting under IMPALA
5. IMPALA Publication Guidelines
6. IMPALA Data sharing, Access and Release Policy
7. IMPALA Data Management Guidelines
8. IMPALA Communications Plan
9. IMPALA kick-off meeting in 2017, annual meetings in 2018 and 2019, including
  1. Meeting schedule
  2. Attendees list and biographies
  3. Meeting slides
10. IMPALA technical reports, including
  1. IMPALA 2017 report (covering the first six months of IMPALA)
  2. IMPALA 2018 annual technical report
  3. IMPALA 2019 annual technical report
11. Research ethics application documents of the two case study projects, including research proposals and data collection tools
12. Quarterly updates by the four post-doctoral researchers each on the two post-doctoral researchers led projects (August 2018 - December 2019, 20 documented updates in total)
13. IMPALA Year 1-3 Joint Outputs list

### Research results

Ten themes (7 themes concerning the *Planning and Implementation* Phases; 3 concerning *Leadership and Management*) emerged from the findings. Interviewee’s anonymised quotes are presented with their main role (researcher/non-researcher) and location (Africa/non-Africa).

#### Actions that fostered CDR in the Planning Phase

##### Shared vision and goals

Interviewees identified that co-development of the IMPALA proposal between members from the Global South and North helped them reach a common vision. While this was time-consuming due to the large number of cross-disciplinary, inter-organisational, and geographically distanced members, several factors helped the process including the existence of previous/ongoing collaborations and involvement in professional associations.

During the face-to-face start-up meeting, IMPALA members and the 22 participating institutions introduced themselves and IMPALA’s vision and strategic objectives were discussed. Specific goals for projects —and for IMPALA as a whole— had purposefully been left undefined by the management team so they could be co-developed during this meeting. Interviewees reported finding this meeting useful for grasping the ‘bigger picture’ of IMPALA, and for learning about one other.

##### Expectations of programme-level goals and success

Interviewees had different expectations of IMPALA depending on their seniority and disciplinary background. Senior, clinical researchers tended to focus on the need to expand collaborations with partners. Two interviewees suggested that since many senior researchers had clinical sciences backgrounds, IMPALA provided more opportunities for clinical researchers to expand collaborations, compared to other programmes. Interviewees from non-clinical disciplines (e.g., social sciences and health systems) were more focused on their existing projects and research quality. Senior researchers sought to enhance ECR’s research skills, and ECRs were focussed on generating outputs and building working relationships. Non-researchers focussed on programme delivery, and capacity strengthening in areas such as financial management, leadership, and policy engagement. All interviewees reported expecting IMPALA to lead to new research questions and new funding. Observation data confirmed all these findings.

Interviewees recognised the complexity of aligning project and programme goals. Two interviewees acknowledged the difficulty of collective prioritisation, and proposed mapping the connections between programme and project objectives, possibly annually. One participant stated that although seeking clarity around programme goals can facilitate members’ engagement, balancing partnership development against addressing a large-scale broad research question with multiple disciplines is difficult.

##### Shared understanding of research questions and activities at the project level

IMPALA’s proposal outlined broad topics for research projects —with named leads and partners for each— while leaving specific research questions and activities to be developed during the start-up meeting. Project leads recognised that this allowed research questions to be based on the interests and experience of partners, and some expressed appreciation that programme leaders had not imposed personal priorities.

Researchers from both Global North and South were comfortable with this process, with Global South partners feeling they had driven the research agenda:

> *“I was looking at ways how I can also contribute rather than just passively engage in national meetings … we were there to conceptualise*…*what we want to do, … we got the research budget.” (ID-21, researcher, Africa)*

Others noted a risk of mismatch between programme and project goals, and had difficulty narrowing research questions down from programme to project level:

> *“When you have multiple perspectives that lead to such a broad potential for research questions that narrowing down and getting in some consensus can be quite difficult.” (ID-1, researcher, Africa)*

The two project teams addressed this differently:

One developed research questions based on a baseline assessment conducted during joint field trips with local research and implementation teams, enabling them to develop locally important, high-priority research questions. To address these questions, they drew on methods from their two core disciplines, indicating some complementary in their disciplinary paradigms such as theories, research methods, and standards. The benefits of having one project integrating two disciplinary components appeared clear to this team from the outset.

The other project team initially generated their research questions independently within each of their two disciplines and then merged the projects through discussions and negotiation which was “*initially uncomfortable*” (*ID-13, researcher, non-Africa*). One researcher believed “*practical efficiency in terms of time and data collection*” (ID-9, researcher, Africa) of this approach to have been the main advantage of merging the two disciplinary research projects into one.

##### Reasons for using CDR in IMPALA

IMPALA took a CDR approach as it was felt its broad research question – i.e. to *address lung health and tuberculosis in Africa* – required inputs from multiple disciplines and programme leaders recognised that everyone had a role in ensuring research findings informed policy. Interviewees considered CDR as one of the “*most effective ways to generate the best possible outputs and outcomes*” (*ID-13, researcher, non-Africa*) since it “*enables appropriate generalisation of research outcomes*” (*ID-15, researcher, non-Africa*). Several interviewees mentioned that multidisciplinary research was a funder’s requirement; however one cautioned “*don’t just do* [CDR] *for the sake of it*” (*ID-14, researcher, non-Africa*).

While most senior researchers recognised the importance of CDR, most interviewees (researchers and non-researchers) had not participated in explicit discussions on what actions would be needed to conduct CDR.

> *“A lot of the challenges is people are so busy doing their own things that they forget that that is what needs to happen.” (ID-12, researcher, non-Africa)*

The IMPALA programme included a post-doctoral researcher (YD) dedicated to investigating cross-disciplinary working. The definitions of multi-, inter- and cross-disciplinary research were presented to IMPALA members during the second IMPALA annual meeting prompting discussions and clarifications. However, interview findings suggest such clarifications would have been useful earlier, alongside discussions on pre-specified goals/methodologies concerning cross-disciplinary working.

##### Cross-disciplinary orientation

Observations clearly indicated that IMPALA members valued understanding more about each other and their disciplines especially within a group environment of psychological safety, while highlighting the value of clarifying disciplinary boundaries to prevent conflicts.

Having inputs from colleagues with various disciplinary backgrounds at the planning phase and arranging formal time for candid conversations on research questions and design were viewed by interviewees as critical. A programme leader and a researcher highlighted potential tensions in cross-disciplinary working, and the need for maintaining ‘*discipline uniqueness*’. The process of defining and clarifying research goals among disciplines was considered to have helped clarify disciplinary boundaries:

> *“After the goals are fixed and then each goal somehow belongs to certain disciplines*…*relate data to that goal and then deal with the data, publication, all those things followed.” (ID-15, researcher, non-Africa)*

#### Actions that fostered CDR in the Implementation Phase

##### Shared understanding of roles and responsibilities

Collaborative working was facilitated by a shared understanding of the roles and contributions of different disciplines and partners, along with an appreciation that successful cross-disciplinary collaborations require complementarity rather than competition. This helped team members to overcome ‘*fighting for space*’ and ‘*struggling for context leadership*’ (ID-22, researcher, Africa). Several interviewees noted the importance of research administrators in helping to understand responsibilities.

> *“Because we [administrators] are that sort of hub in the middle, and we do oversee everything. We can sort of speak on behalf of the project and say that this isn’t working and have a bit of input in that way.” (ID-3, non-researcher, non-Africa)*

Several interviewees had not had open discussions about roles and responsibilities, with one suggesting that roles were defined by one’s job description and another explaining that “*as a member of the team you naturally know your strengths and therefore role*” (ID-5, researcher, Africa). Another interviewee highlighted that assumptions regarding roles and responsibilities had the potential to cause confusion and needed open discussions:

> *“I increasingly think the best way to have good, harmonious, collaborative relationships is to be really upfront about roles and responsibilities. To do that first so that there is no confusion after.” (ID-9, researcher, Africa)*

One interviewee suggested that jointly developing a work plan, containing explanations of responsibilities alongside a clear timeline could help to clarify roles.

##### Reconciling individual expectations while navigating different contexts

Several interviewees advocated for open discussions on roles, suggesting such discussions were important because people were at different career stages with different experiences, cultures and academic systems, which could cause mismatched expectations of one another’s roles. This led to, for example, disagreements on the time spent in the research sites and responsibilities for research coordination. Clarifying roles and having a host country/institution coordinator was thought to be essential in avoiding these issues.

Regular cross-disciplinary project update meetings along with individual conversations to provide performance feedback to ECRs (including those with different disciplinary backgrounds) were said to be useful by both ECRs and senior researchers. Role modelling was also identified as important in encouraging ECRs to continuously explore other disciplines:

> *“Seniors and line managers say, ‘You should go to this. Think about this*…*’ So, it does need people, at a senior level, to think broadly and encourage that.” (ID-23, researcher, non-Africa)*

Support across disciplines was valued during project implementation, for example when developing questionnaires, collecting and analysing data and several senior researchers called for more thought on how to provide supportive supervision:

> *“Perhaps we didn’t think hard enough about how to support the projects and who should be supporting the projects and in what way.” (ID-9, researcher, Africa)*

##### Team learning

The importance of individuals’ ability to blend disciplinary edges was raised by an interviewee and many others shared their approaches to understanding other disciplines. Senior researchers also encouraged colleagues to consider broadening the scope of their work and skillset through formal cross-disciplinary training, mutual learning, and joint supervision in other subject areas. One month after the interviews, monthly knowledge exchange meetings were initiated to improve cross-disciplinary learning and communication.

#### Leadership and Management

##### Communication planning and implementation

New IMPALA members appreciated their one-to-one induction meetings with key researchers and administrators. Joint site visits by members from the Global North and South were helpful in forming relationships and in promoting cross-fertilisation. Face-to-face meetings were valued for facilitating the design, prioritisation, and development of both research projects and teams, especially concerning developing methods and budgeting. Interviewees said that virtual meetings and email communications worked well and were useful, though several raised issues with internet connections. Effective planning to maximise the availability of team members was highlighted:

> *“What I usually do is to inform them early enough because they have lots of responsibilities…After they have considered then you block the time*… *With multi-disciplinary, it needs proper planning, especially on timing.” (ID-26, researcher, Africa)*

Many senior researchers often had long working relationships with country partners. To help ECRs to build mutual understanding and develop research networks, regional meetings for ECRs across disciplines were suggested.

Several interviewees suggested that having access to other teams’ materials and outputs could have improved cross-disciplinary understanding. A common platform for document and information sharing was subsequently established. Interviewees further proposed that cross-disciplinary communications should be expanded. Accordingly, the monthly knowledge exchange meetings were expanded beyond ECRs to include administrative staff, in-country partners, and researchers beyond IMPALA’s core team.

Interviewees wanted more time to develop mutual understanding in CDR and to create a sense of ownership. One interviewee reflected “*we need to have some more recognition of the need for time for some of the processes and the collaborations to work for the future*” (*ID-11, researcher, non-Africa*). Another recommended taking time to learn about each other’s experiences and expectations, ways to successfully collaborate, and for joint preparation of project tools (e.g. databases).

According to several interviewees “*there are inevitable delays in starting*” (*ID-9, researcher, Africa*) *for example* in funding release (6 months), international staff recruitment (5-8 months) and ethics approval (7-8 months). Interviewees described how they felt the need to focus on outputs although “*would have loved to have used those six months to think about how we prepare these disciplines to work together*” (*ID-11, researcher, non-Africa*). One interviewee highlighted the importance of prioritising internal communication even within tight timescales, arguing “*sometimes prioritising a two-hour meeting to make sure everyone’s on the same page and understanding things in the same way is equally important as papers and research outputs*” (*ID-11, researcher, non-Africa*).

##### Nurturing trust and a group environment of psychological safety

Two senior researchers, three ECRs and three non-researchers noted that IMPALA management had helpfully promoted involvement and empowerment of ECRs and non-researchers and two ECRs appreciated the space and freedom their line managers had given them to lead projects.

There were three other suggestions offered by interviewees for nurturing trust:

i) Treating everyone equally through “flat management”:

> *“I very strongly believe in flat management, a structure everybody is equal. If I have a research meeting in my team, they all know we are equal. If they have something to say, they are all happy to say, and confident to say it.” (ID-2, researcher, non-Africa)*
ii) Building trust by delivering on commitments (mentioned by two researchers and one non-researcher):

> *“To build trust you need to deliver*… *I think that’s important, showing that you want to do your best. Then by reflection they don’t want to let me down, so they deliver, and that’s how you build trust, I think.” (ID-4, non-researcher, non-Africa)*
iii) Being transparent and learning from mistakes.

> *“Transparent, I think building trust… Also within trust and team, you have to allow mistakes… Accepting and also sitting together and see how we can handle it next time.” (ID-21, researcher, Africa)*

##### Addressing disciplinary hierarchies through the management structure

According to three interviewees, disciplinary hierarchies emerged when one discipline’s work depended on another. For example, when one discipline’s research questions and analysis relied on another’s data generation, the latter may perceive their research activities should be prioritised over the former. Despite both projects having been allocated equivalent resources at the start of IMPALA, perceived imbalances arose. Five interviewees suggested that since clinical aspects were the primary interest of several IMPALA leaders, this may have inadvertently contributed to disciplinary hierarchies. Furthermore, several interviewees found the equal allocation of resources limiting, potentially hindering the effective answering of some research questions. Two interviewees further noted that since studies were highly interconnected at the operational level, strict drawing of financial boundaries between projects could at times “*lead to tensions*” (*ID-1, researcher, Africa*).

Following the initial equal allocation of resources, a degree of re-negotiation continued throughout IMPALA’s lifetime though some members questioned the success of this process. One remarked that “*an alternative approach may be to develop the budget based on justified activities*” (*ID-15, researcher, non-Africa*).

##### Handling disciplinary differences and managing emotions

At times, the different approaches and priorities of disciplines led to some disagreements. Overall, the group which combined *Clinical Sciences* and *Health Economics* was perceived as predominantly outputs-driven whereas the *Humanities and Social Sciences group* appeared primarily focussed on processes, consultation, and discussions. One interviewee from the Management Team reflected “*we probably hadn’t paid enough attention to the need for the process [of discussions between the management team members]*” because it “*requires sustained effort to balance the natural priority of an individual’s discipline against that of multiple disciplines*” (*ID-11, researcher, non-Africa*). Two interviewees suggested that time spent discussing managerial and logistics issues could have been more productively spent on research activities and constructive management of disciplinary disagreements.

Several interviewees described encountering emotional challenges, most frequently caused by disciplinary differences and some identified having needed for dedicated meetings to manage emotions in a professional environment. One interviewee commented that their previous working relationships and sense of responsibility had helped to make these conversations possible.

Such conversations resulted in real-time adaptations to the programme to enhance cross-disciplinary relationships. For example, monthly Directorate and Management Team meetings were merged and a rotational system for the management meeting chair was instigated whereby each discipline lead and the consortium directors took turns in chairing. Handovers between meetings were supported by the Programme Management and Administration support staff. Actions to promote more effective cross-disciplinary collaborations were also identified through a group exercise during the second annual meeting, and reviewing uptake of these actions became a standing item at management meetings. These changes were viewed as positive by several interviewees.

##### Developing research networks

Interviewees emphasised the programme’s many good working relationships between different partners across Global North and South and noted the considerable benefit from strong previous relationships of key leaders. The importance of enabling the development of such research networks was a repeated theme from interviews.

> *“I think it [IMPALA] has really done a great job bringing great collaborators in terms of Africa with Liverpool, countries that are involved. I think it’s really an interesting network and it has brought us together, many collaborators. People have never even met.” (ID-21, researcher, Africa)*

Two interviewees reflected that project activities had helped build up trust and develop research networks.

> *“I hope my work […] will let them [current IMPALA members] say that ‘he would actually put the neck on the line and physically help you. Get him on board.’” (ID-2, researcher, non-Africa)*

##### Strengthening capacity

Several approaches to capacity strengthening were identified through interviews and corroborated by internal documents. These included:

- Training workshops for those with different disciplinary backgrounds from the training subjects (e.g., training on social science research methodologies, policy engagement, statistics, and spirometry)
- Coaching through team meetings and one-to-one discussions (e.g., two interviewees emphasised that discussions with a statistician catalysed research)
- Mentoring ECRs and providing them with platforms at high-level international meetings (e.g., the UN General Assembly)
- Learning through peer support and reflection was mentioned by ECRs, senior researchers, and non-researchers:

> *“I feel like I’m definitely learning a lot… It’s nice working so closely with […] and she’s able to delegate things to me as and when they come up.” (ID-3, non-researcher, non-Africa)*

Capacity strengthening also involved administration and field teams:

> *“My ideal world would be a world where everyone can do it because that’s capacity building in-country. And it is not just the research, it’s the admins.” (ID-4, non-researcher, non-Africa)*

## DISCUSSION

We adapted and expanded a published framework to underpin our research. Our findings emphasise that CDR programmes require careful planning, implementing, and managing and we have identified actions to promote CDR including some that have not previously been published.

### Actions in programme planning to foster CDR

#### Clarity in defining ‘Cross-disciplinary Research’

Similar to other studies we found a lack of agreement on defining multi-, inter- and trans-disciplinary research.[16] Our findings demonstrate that explicit discussions concerning both these definitions and what CDR means in practice are critical in the Planning Phase.

#### Managing expectations and harmonising goals

Participants had different expectations about being involved in CDR and highlighted the importance of negotiating a clear shared vision, taking into consideration individuals’ expectations.[17] To harmonise goals, frequent discussions and interactions such as information sharing can be helpful, [3, 18] and need to be more frequent and intensive than in mono-discipline research.[9] Our findings shed light on tensions that can arise early in CDR, including balancing flexibility and acceptance that not all aspects of the research could be initially ‘nailed down’, with developing a common understanding of the goals.

As with previous studies, IMPALA’s participants recognised the importance of a common conceptual framework for outlining the vision, objectives and organisational structure for showing the contributions of each discipline,[19] and to guide collaborations.[17, 20] Furthermore, evidence suggests that to have explicit knowledge integration goals for CDR is helpful.[20, 21] IMPALA’s conceptual framework was strengthened during the programme, for example by taking account of local contexts (achieved through joint field trips and discussions), by co-developing research questions and by drawing methods from relevant disciplines.

#### Actions in programme implementation to foster CDR

Our findings reflect previous studies which suggest that cross-disciplinary relationships flourish if they are prospectively planned and actively monitored.[3, 9] This is best managed separately from activities that focus on research outputs since fostering cross-disciplinary relationships requires its own planning and activities,[22] specific monitoring indicators and mechanisms for collecting data against the indicators.[23]

### Management actions to foster CDR

#### Development of research collaborations and networks

Our study revealed important findings concerning management strategies for encouraging equitable partnerships, fostering CDR and reconciling individual expectations. These included involving northern and southern partners in co-developing a shared vision and goals, designing project-level research questions and activities, and strengthening capacity in line with a baseline capacity assessment.

#### Allowing time to promote cross-disciplinary activities

Our research also identified that researchers lacked sufficient time to successfully engage in discussions and processes to promote cross-disciplinary activities. Building in adequate time and funds for this throughout the programme is critical and may necessitate a shift in research planning as well as an understanding among research funders that such allocations are essential.

Lack of time for active consideration and management of activities to promote cross-disciplinary working is closely linked to lack of effective communication among programme members to bridge across disciplines.[24, 25] Interviewees proposed that cross-disciplinary communications should include all team members. This requires an agreed internal communication plan, administrative support and an electronic communication platform. Other studies have also highlighted the importance of an accessible space to document programme work and decision making.[26]

#### Programme adaptations to address hierarchies and tensions

Our framework specifically recognised ‘nurturing trust and a group environment of psychological safety’, ‘communication planning and implementation’, and ‘team learning’ in CDR as important because of possible emotional issues associated with ownership, territoriality, academic and discipline hierarchy, and disciplinary differences. Similar to previous studies, our findings identified CDR-related emotional issues (particularly around power and hierarchy) and disagreements in disciplinary approaches.[17, 27, 28] IMPALA took measure to mitigate such frictions, including providing equal funding and training opportunities, and by adjusting the programme’s management structure. There included merging monthly Directorate and Management Team meetings, having a rotating Chair for Management Team meetings, initiating monthly knowledge exchange meetings for mutual learning and cross-disciplinary communications, and creating a common platform for document and information sharing. In addition, emerging findings from our study on CDR were presented at Management Team meetings and summarised in quarterly bulletins for all IMPALA members so that they could inform subsequent programmes.

### Strengths and limitations of the study

As members of IMPALA our research team generally had easy access to programme colleagues and documents, as well as opportunities for informal discussions, though we were not involved in decision-making at the programme and project levels. Our findings throughout the project — especially when deemed of benefit for enhancing cross-disciplinary working—were fed back at Management Team meetings. We were aware that our role in conducting research on cross-disciplinary working in IMPALA may affect interviewees’ responses. To mitigate our influence, we ensured that our interviewees understood that their participation was voluntary, that data collected would be handled confidentially, and that our findings would be reported anonymously. The internal validity of our findings is strengthened from having used interview and observational data from diverse interviewees and events, corroborated by document analysis.[29] Although our study focused on a single cross-disciplinary global health research programme and its projects, we have enhanced the generalisability of our findings by describing the complexity of the programme and the context within which the CDR took place. We used an adapted published framework and a recent literature review to frame our data collection tools and analysis, and have placed our findings in the context of current global knowledge concerning CDR.

### The three-component framework on CDR

Using our ‘real-life’ findings we adapted and expanded a published model of cross-disciplinary collaborative research processes[9] to create a framework useful for collecting and analysing multi-source and multi-perspective data on CDR in real-time. A new component of the framework emphasised the importance of leadership and management in CDR processes. We would recommend further adaptations to the framework to include a rationale for the components and to expand the ‘shared understanding of who knows what, who does what, and how things get done’[9]. In addition to being useful for future research on CDR, our framework could be used to guide the design of cross-disciplinary programmes since it has practical applications across the three programme components of planning, implementation and management (Figure 2).

### Recommendations

Based on the findings from our study, our adapted framework and our knowledge of the current literature, we have developed recommendations for planning and implementing future CDR in global health to improve the effectiveness of CDR processes from the outset (Table 2).

**Table 2:** Recommendations for the planning, implementation and management of cross-disciplinary global health research

| Research Phase | Recommendations |
| --- | --- |
| Planning Phase | 1) Allocate adequate time to develop a shared vision and goals, including<br>a. Co-designing of programme goals;<br>b. Aligning individuals’ expectations and projects’ aims with the programme-level goals;<br>c. Involving all partners in proposal development, maintaining flexibility, considering individual interests and disciplines;<br>d. Justifying and communicating the cross-disciplinary approaches to be adopted and reflecting cross-disciplinary processes in an action plan;<br>e. Developing and communicating a shared understanding of the roles, responsibilities and potential contribution of disciplines and partners.<br>2) Negotiate disciplinary boundaries when necessary;<br>3) Assess and strengthen in-country teams’ capacity in cross-disciplinary research, and maintain clear plans for the involvement of in-country |
|  | teams in decision making processes. |
| Implementation Phase | 1) Jointly develop and pre-agree on internal approaches of working across disciplines, including communication, data access and management, publication policy and credit allocation;<br>2) Track the implementation of cross-disciplinary processes with pre-agreed indicators and review and respond accordingly. |
| Leadership and management | 1) Rotate chairs for programme management meetings to ensure prominence of all relevant disciplines and with a process for handover and preparation between meetings.<br>2) Define and agree transparent programme-level mechanisms for strategic decision making;<br>3) Develop a programme-level leadership and management plan to deliver the cross-disciplinary outputs and outcomes, including regular review of tracking indicators;<br>4) Agree roles and responsibilities, and accountabilities, and communicate these clearly to all programme members, making it explicit that every role is important in cross-disciplinary research (i.e. not just researchers).<br>5) Support an open culture of raising concerns and putting mechanisms in place for requesting support and responding to requests |
|  | 6) Establish mechanisms for early identification of tensions, and for reflecting on and flexibly resolving differences and conflicts<br>7) Provide opportunities for joint learning and knowledge exchange across disciplinary boundaries especially methods and approaches (e.g., monthly knowledge exchange meetings)<br>8) Identify a platform for joint sharing and updating of documents. |

## Data Availability

The present study includes selected quotes to represent the content and themes across interviews.  While requests for additional de-identified transcripts can be made to the authors, full interview transcripts cannot be made available as to do so would compromise anonymity and violate the terms of our ethical approval.  

## Acknowledgements

We are grateful to all participants who agreed to take part in this case study and provided their valuable information and insight. We are also thankful to the IMPALA central administration team and the Centre for Capacity Research administration team, Annmarie Hand, Elly Wallis, Lorelei Silvester, Zena Parker who made the necessary logistical or administrative arrangements to allow a smooth data collection process. We would like to thank our Research Impact & Knowledge Translation Officer, Susie Crossman for her support in reviewing and editing the manuscript. We are grateful to Martina Savio, Dr Angela Obasi and Professor Stephen Bertel Squire from the IMPALA Management Team for their review of an earlier draft.

This research was funded by the National Institute for Health Research (NIHR) (project reference 16/136/35) using UK aid from the UK Government to support global health research. The views expressed in this publication are those of the author(s) and not necessarily those of the NIHR or the UK Department of Health and Social Care.

## Contributors

YD: Conceptualization, Methodology, Data Curation, Writing-Original draft preparation

EW: Methodology, Writing-Reviewing and Editing.

IB: Conceptualization, Methodology, Writing-Reviewing and Editing, Supervision.

All authors read and approved the final manuscript.

